# Aromatic L-Amino Acid Decarboxylase is a novel fluid biomarker of Parkinson’s disease

**DOI:** 10.1101/2022.11.09.22282149

**Authors:** Jarod Rutledge, Benoit Lehallier, Pardis Zarifkar, Patricia Moran Losada, Sephira Ryman, Maya Yutsis, Gayle Deutsch, Elizabeth Mormino, Alexandra Trelle, Anthony D Wagner, Geoffrey Kerchner, Lu Tian, Victor W. Henderson, Thomas J Montine, Per Borghammer, Tony Wyss-Coray, Kathleen L Poston

## Abstract

There are currently limited molecular markers of Parkinson’s disease, and there is an urgent need for new markers to guide clinical care, support earlier diagnosis, and hasten drug development. Here, we performed CSF and plasma proteomics in 5 Parkinson’s disease cohorts to identify novel protein biomarkers for these purposes, resulting in one of the largest such resources for Parkinson’s disease to date. We discovered a consistent upregulation of the protein L-Aromatic Acid Decarboxylase (AADC, EC 4.1.1.28, DDC) in the CSF and plasma of Parkinson’s disease patients. AADC is a key protein in the synthesis of dopamine and other monoamine neurotransmitters. We found that higher CSF AADC levels are associated with greater motor symptom severity in Parkinson’s patients. We replicated and extended these findings in another undescribed proteomics cohort of *de novo* Parkinson’s disease participants from the Parkinson’s Progression Marker Initiative, where we found that AADC expression is upregulated in treatment naïve participants and is associated with motor and cognitive symptoms. We found that AADC expression can accurately distinguish Parkinson’s disease from healthy participants and Alzheimer’s disease participants in multiple independent cohorts, and developed a panel of 16 proteins that achieves 95% receiver operator area under the curve (ROC AUC) in distinguishing these three states. Our results suggest that CSF AADC is a marker of the underlying disease process in Parkinson’s disease with potential utility in multiple contexts.

## Introduction

Parkinson’s disease (PD) is characterized by selective neuronal degeneration of the substantia nigra and the accumulation of alpha-synuclein protein aggregates in the brain^1^. The defining histopathologic feature of the disease is the loss of dopaminergic neurons in the substantia nigra but can also include noradrenergic neurons in the locus coeruleus^2^ and serotonergic neurons in the dorsal raphe nuclei^3^. Notably, there is a delay between the first damage to the dopaminergic neurons and the development of clinical motor symptoms, which makes diagnosing PD in the earliest stages of neurodegeneration very challenging^4^. Among those at risk for PD it is even more difficult to identify who has already developed asymptomatic neurodegeneration. There is an urgent need for biomarkers to diagnose PD early and accurately, and to assess disease severity, progression, and response to therapeutics^5–7^. Biomarkers that reflect the earlier, underlying pathophysiologic processes or that reflect early compensatory mechanisms that preclude symptom development despite active neurodegeneration hold most promise for disease identification before clinical symptoms develop. The power of cerebral spinal fluid (CSF) biomarkers that are specific for the signature pathology underlying neurodegeneration, which can aid in early diagnosis and therapeutics development, has been convincingly shown by the Alzheimer’s disease (AD) field. Beta amyloid (Aβ) and Tau biomarkers can be identified as abnormal in individual patients prior to cognitive decline^8^, and have revolutionized translational AD research over the last decade; however, there are no analogous biomarkers for PD.

Quantitative proteomics has recently been used to develop disease-specific protein signatures as diagnostic biomarkers and holds great promise to enhance our current understanding of the molecular mechanisms underlying neurological diseases. In this study, we quantified the levels of 1,196 proteins in CSF and blood plasma in five human neurodegenerative disease cohorts to identify disease-specific protein signatures of PD that correlated with observed disease severity and could distinguish PD participants from cognitively normal individuals and Alzheimer’s disease participants.

## Methods

### Participants

We included participants from five concurrent longitudinal cohorts of aging and neurodegeneration at Stanford University: 1) Biomarkers in PD Study (BPD), 2) the Pacific Udall Center (PUC), 3) Stanford Alzheimer’s Disease Research Center (ADRC), 4) Stanford Center for Memory Disorders Cohort Study (SCMD), and 5) Stanford Aging and Memory Study (SAMS). Data were collected between 2012 and 2018. Inclusion criteria for these analyses were i) ages between 40 and 90 years ii) English or Spanish fluency for comprehensive neuropsychological testing, and iii) no contraindications to lumbar puncture. All participants provided written informed consent to participate in the parent studies following protocols approved by the Stanford Institutional Review Board.

A consensus panel consisting of one board-certified movement disorders neurologist or behavioral neurologist, one board-certified neuropsychologist, and other study personnel adjudicated the diagnosis for each participant. PD diagnosis was based on UK PD Society Brain Bank clinical diagnostic criteria^9^. We defined Early PD as participants with less than three years disease duration at the time of CSF collection. Participants on the AD spectrum (AD-s) included those with dementia or mild cognitive impairment likely due to AD based on the NIH Alzheimer’s Disease Diagnostic Guidelines^10,11^. Participants with mild cognitive impairment, who have decreased CSF Aβ-42 concentration, are more likely to have cognitive impairments due to AD^12^. To exclude participants without AD from the AD-s group, we excluded mild cognitive impairment participants who had CSF Aβ-42 concentration more than 2 standard deviations from the mean in AD^13,14^. Participants with PD include those with no cognitive impairment, those with mild cognitive impairment^15^ and those with dementia due to PD. Healthy controls (HC) were older individuals without a neurological diagnosis adjudicated as cognitively normal for age at the consensus meeting. A flowchart of included and excluded participants is illustrated in extended data figure 1.

### Neurologic, Motor and Cognitive assessments

All participants completed a general neurological exam. PD participants completed the Movement Disorders Society-Unified Parkinson’s Disease Rating Scale (MDS-UPDRS III)^16^ in the Off- and On-medication states, according to published criteria^17^. We calculated the Levodopa Equivalent Daily Dose (LEDD) using previously reported conversion factors^18,19^.

Global cognitive function was assessed using the Montreal Cognitive Assessment (MoCA)^20^ in the ADRC, PUC and BPD, and the Mini-Mental State Exam^21^ in the SAMS and SCMD studies. A comprehensive neuropsychological battery was administered as part of each cohort study as previously described^22–25^. PD participants underwent neuropsychological testing in the on-medication state in order to assess cognitive function without interference by motor deficits.

### CSF collection and assessment

A neurologist performed a lumbar puncture to collect CSF samples according to standardized procedures^26^. Briefly, a 20-22 G spinal needle was inserted in the L4-L5 or L5-S1 interspace and CSF was collected in polypropylene tubes. The tubes were immediately frozen at −80 °C in a centralized freezer in the Neuropathology Core of the Stanford ADRC and sent to Quanterix (Quanterix ®, MA, USA), for the quantification of seven major neurology biomarkers present in the Neurology 3-plex A assay (total Tau, Αβ42, Aβ40), Neurology 4-plex A assay (Nf-L, total Tau, GFAP, UCHL1) and p-Tau181.

### CSF and plasma proteomics

201 CSF and 249 blood plasma samples, (including 173 matched samples from the same participants) were sent to Olink Proteomics AB (Uppsala, Sweden. www.olink.com) for the quantification of 1196 proteins in each tissue using a multiplex proximity extension assay^27^. This technology has been extensively vetted in biomarker studies and detailed methodology of the assay has been previously published^27^. Briefly, the proximity extension assay uses DNA oligonucleotide-labeled polyclonal antibodies which bind to each protein target. When two antibodies targeting different epitopes bind the same protein target, a proximity-dependent DNA ligation and elongation reaction can occur. The requirement for coincident binding leads to high specificity. The target protein levels can then be read out using quantitative PCR (qPCR). This technology enables multiplex measurement of 96 protein targets in a single assay. Proteins from 13 different protein panels were measured, resulting in quantification of 1,196 CSF and plasma proteins. CSF and plasma protein levels were analyzed using the Cardiometabolic (v.3602), Cardiovascular II (v.5005), Cardiovascular III (v. 6112), Cell Regulation (v.3701), Development (v.3512), Immune Response (v.3201), Inflammation (v.3012), Metabolism (v.3402), Neuro Exploratory (v.3901), Neurology (v.8011), Oncology II (v.7002), Oncology III (v.4001) and Organ Damage (v.3301) 96-plex immunoassay Olink panels.

### Proteomics data processing and quality control

As previously described^27^, 8 control samples are run on each plate: two are external pooled plasma samples, which are used to assess potential intra-plate/run variation, three are Inter-Plate Controls (IPCs) and three are buffer blanks. The IPCs are formed from a pool of 92 antibodies. The median of the IPCs is used to normalize each assay and compensate for potential variation between runs and plates.

Protein expression data are reported in Normalized Protein eXpression (NPX), which is a normalized unit on a log_2_-scale. The NPX values are derived from the Ct or “threshold cycle”. This is the number of qPCR cycles needed for the signal to pass a fluorescence signal threshold. NPX is calculated from the Ct values using the following equations:

Extension Control: *CtAnalyte – CtExtension Control = dCtAnalyte*

Inter-plate Control: *dCtAnalyte – dCtInter-plate Control = ddCtAnalyte*

Adjustment against a correction factor: *Correction factor – ddCtAnalyte = NPXAnalyte*

### Statistical Analyses

To examine demographic and clinical group differences, we used an independent sample t-test or a one-way analysis of variance (ANOVA) for normally distributed variables, or a non-parametric Wilcoxon sign-rank test or a Kruskal Wallis H-test for non-normally distributed variables. We performed post-hoc Tukey correction for multiple comparisons.

We ran differential expression analysis on protein levels using a multi-level linear-mixed effects model controlling for age, sex, race, ethnicity, and sample-relatedness when using longitudinal sample data. Sample-relatedness refers to longitudinally collected samples from a single individual, which we expect to be more correlated than samples from different individuals. We used a linear model controlling for age, sex, race, and ethnicity when looking at samples from one timepoint only. We used Benjemani-Hochberg false discovery rate control to account for multiple testing. We studied the association between AADC levels and clinical measures of disease severity (MDS-UPDRS III, LEDD, MoCA) using linear regression analyses corrected for age, sex and education. We used principal component analysis to explore the relationship between global differences in protein expression profile and clinical/demographic variables. We tested correlations between principal components with a spearman correlation test with Bonferroni correction for multiple testing. All statistical analysis was done in R 4.0. We used the package lmerTest^28^ and the dream function from the R package variancePartition^29^ for mixed effects models.

### Disease classification

We used the glm function with a binomial link in R 4.0 to perform binary logistic regression. We used the step function in R 4.0 to perform step-wise logistic regression when building multivariate linear classifiers, and used the Akaike Information Criterion for model selection. We used the multinom function from the R package nnet^30^ with L2 regularization for multi-class logistic regression. We used the caret package^31^ to perform cross-validation (CV) on our multiclass models, which was used to optimize regularization and estimate model performance. We used a repeated 2×10 CV scheme to minimize overfitting: each model instance in the CV was trained on half of the data and evaluated on the other half. We used the pROC^32^ and multiROC^33^ packages in R 4.0 to generate and visualize receiver operator sensitivity-specificity curves and calculate area under the ROC curve.

## Results

### Participant characteristics

Table 1 lists demographic and clinical characteristics of the CSF study population. The final cohort included 71 PD, 78 HC and 52 AD-s, who were matched for age and sex but not education (p=0.033). Twenty-one PD participants were early in disease (less than three years duration) and two were dopamine naïve.

**Table 1:** Demographic and clinical participant characteristics of CSF study population. Kruskal Wallis H-test was used to determine group differences in non-normally distributed variables (education, MoCA and MMSE). *MoCA score is lower in AD-s compared to HC and PD. Education is higher in HC compared to AD-s. The table presents mean ± standard deviation (range) with bolded values indicating *p* < 0.05. AD-s, Alzheimer’s Disease spectrum; F, female; HC, Healthy Controls; Levodopa Equivalent Daily Dose, LEDD; M, male; MMSE, Mini-mental state exam; MoCA, Montreal Cognitive Assessment; PD, Parkinson’s Disease.

|  | HC | AD-s | PD | *P*-value |
| --- | --- | --- | --- | --- |
| **N** | 78 | 52 | 71 |  |
| **Age (yrs)** | 67.7 ± 7.4<br>(55-87) | 67.7 ± 8.71<br>(41-85) | 68.6 ± 7.56<br>(50-85) | 0.763 |
| **Education (yrs)** | 17.1 ± 2.0<br>(12-20) | 16.1 ± 2.3<br>(12-20) | 16.0 ± 2.3<br>(12-20) | **0.033\*** |
| **Sex** | 49.5% F, 50.5%<br>M | 48.8% F, 51.2%<br>M | 46.6% F, 53.4%<br>M | 0.950 |
| **MoCA** | 27.2 ± 2.3<br>(19-30, n=48) | 14.6 ± 8.0<br>(1-26, n=14) | 24.4 ± 5.0<br>(8-30) | **<0.001\*** |
| **MMSE** | 29.5 ± 0.9<br>(25-30 n=45) | 26 ± 3.4<br>(17-30, n=26) | -- | **<0.001\*** |
| **MDS-UPDRS III Off** | -- | -- | 36.7 ± 13.0<br>(6-75) | -- |
| **MDS-UPDRS III On** |  |  | 23.6 ± 11.0<br>(4-51) |  |
| **Duration of PD (in years)** | -- | -- | 6.1 ± 4.3<br>(0-21) | -- |
| **LEDD** | -- | -- | 626.9 ± 389.5<br>(0-1580) | -- |

### CSF and plasma proteomics identify biomarkers of PD

We first compared the CSF and plasma proteomes of people with PD or AD-s and HC. Exploratory principal component analysis (PCA) including all CSF and plasma proteins indicated that there were not strong global differences in protein expression between the disease groups or reported gender. The global CSF proteome profile did show a significant correlation between the first principal component and age (spearman p = 0.0005) (Extended data figure 2). The plasma proteome showed no significant global correlation with age.

To identify proteins whose expression differed across groups, we next performed differential protein expression analysis on the CSF and plasma proteomes using linear mixed-effects models while controlling for age, sex, education, ethnicity, and sample relatedness (Figure 1). When comparing people with PD to HC, there was 1 significant hit in CSF (Figure 1A, Supplementary Table 1) and 10 significant hits in plasma (Figure 1B, Supplementary Table 2) after multiple testing correction. Comparing people with PD or AD-s there were 3 significant hits in CSF (Figure 1C, Supplementary Table 3) and 9 significant hits in plasma (Figure 1D, Supplementary Table 4). We also performed differential expression for age and sex (Extended data figure 3, Supplementary Tables 5, 6, 7, 8), and replicated known top hits such as PTN and WFDC2 as significantly up in aging in plasma^34,35^, and CGA and CGB3 as significantly differential between sexes^35,36^.

**Figure 1.**
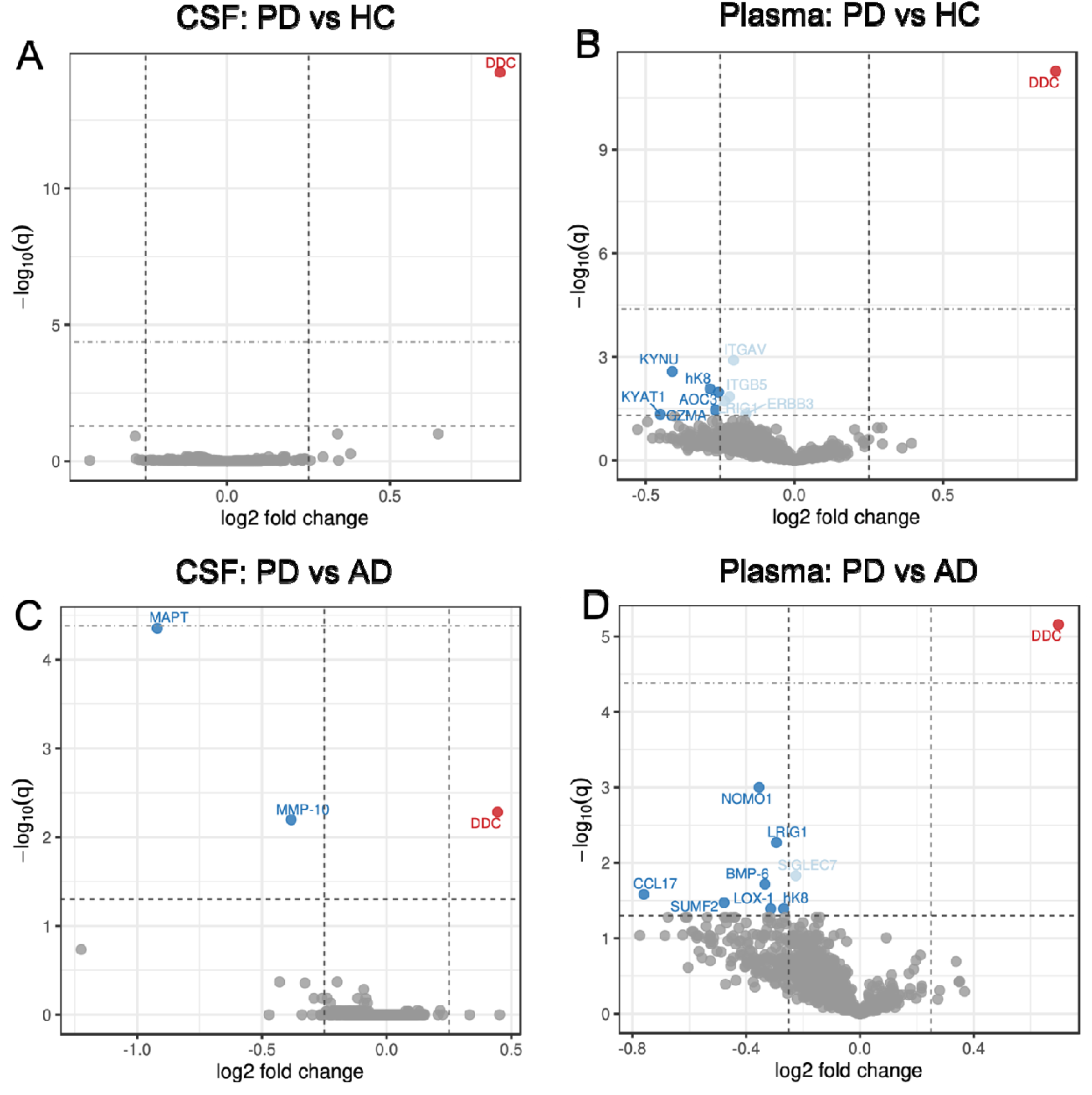
Differential expression analysis of Parkinson’s disease participants in our 5 cohorts using a multi-level mixed effects model controlled for age, sex, demographics, and sample-relatedness. Lower horizontal dotted line indicates FDR significance threshold, upper horizontal line indicates Bonferroni significance threshold. Dotted vertical lines indicate an arbitrary 0.25 log2 fold change cutoff for differential expression. Significant hits are colored and shaded by cutoff. DDC encodes the enzyme AADC, it is labeled here as DDC to remain consistent with the naming convention of the Olink proteomic platform. A). Results comparing PD participants to healthy controls in CSF. B). Results comparing PD participants to AD-s participants in CSF. C.) Results comparing PD participants to healthy controls in plasma D.) Results comparing PD participants to AD-s participants in plasma.

Notably, the protein Aromatic L-Amino Acid Decarboxylase (AADC EC 4.1.1.28), also known as DOPA Decarboxylase (DDC), was the top upregulated hit in PD when compared to both HC and AD-s, in both CSF and plasma. We elected to explore this further because AADC is directly involved in dopamine synthesis in dopaminergic neurons and is highly expressed in the substantia nigra (Extended data figure 4A). This mechanistic link to known PD pathogenesis makes it an appealing biomarker candidate.

### AADC expression is associated with disease symptom severity in five independent cohorts

After identifying AADC as a top hit in the differential expression analysis for PD, we directly compared AADC CSF levels in PD to HC and AD-s. We found a significant difference across groups (F_2_,_207_=47.7, *p*<0.001). Specifically, AADC is elevated in PD compared to both HC and AD-s (Figure 2A). AADC remained significantly elevated after controlling for age, sex and LEDD, (F_2,200_=10.515, *p*<0.001, partial η^2^=0.095). We further validated this finding using an orthogonal proteomics platform, the SomaScan assay^37^, and found CSF AADC to be significantly upregulated in PD participants compared to both AD and HC (Extended data figure 4B). We then studied AADC as an early marker of PD and found significantly elevated concentrations in Early PD (<= 3 years since symptom onset) compared to HC, F_113_=9.101, *p*<0.001 (Figure 2B). We next tested if ADDC levels in CSF and plasma were associated with PD symptom severity and could thus have potential prognostic and clinical utility. We found that AADC levels in CSF were significantly associated with severity of motor symptoms assessed on the MDS-UPDRS III Off (β = 2.85, p = 0.022) (Figure 2C), and On scores (β = 3.29, p = 0.0017) (Figure 2D).

**Figure 2.**
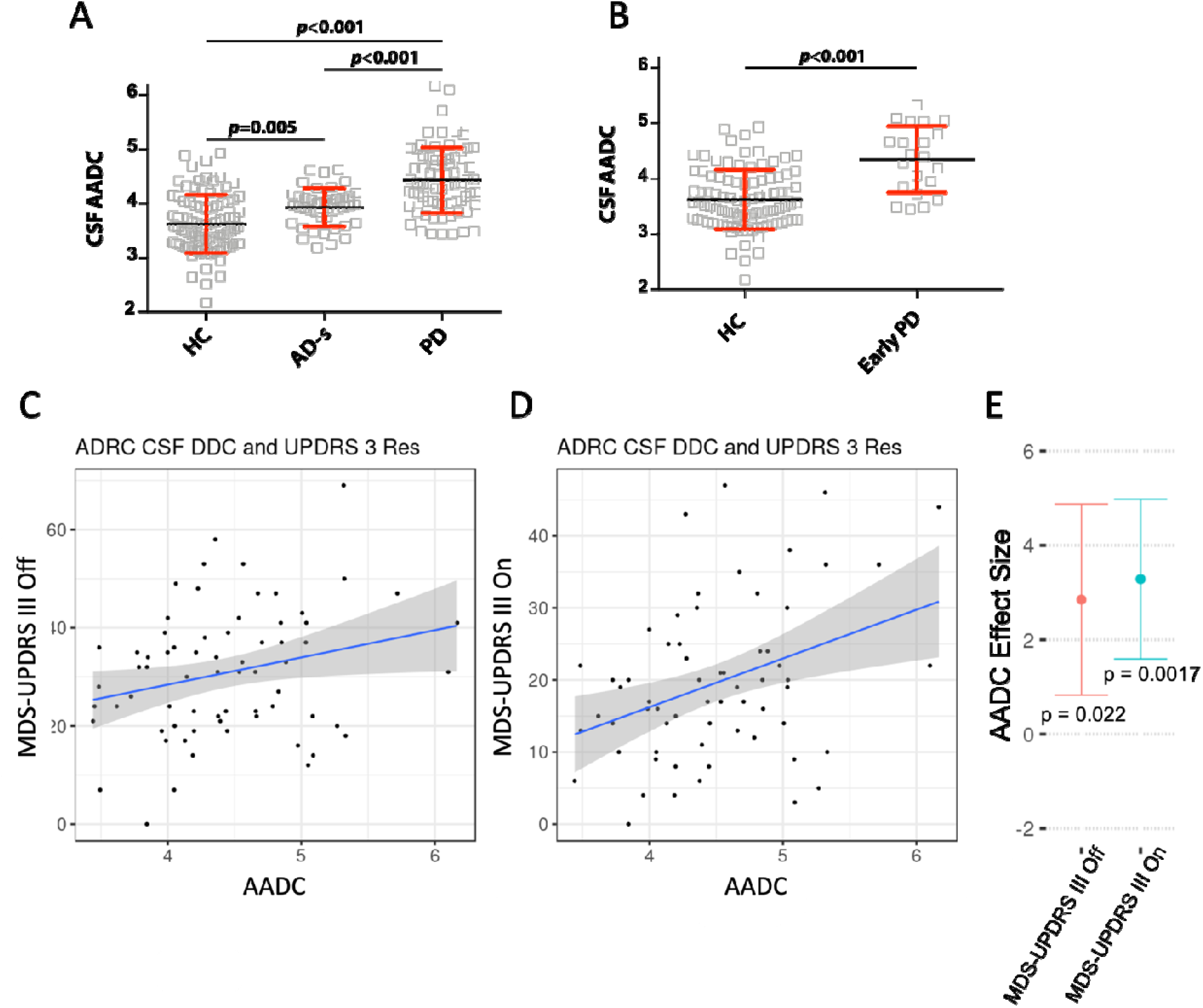
A). CSF AADC levels are elevated in AD and PD participants, with greater elevation in PD. Test statistics from Wilcoxen Rank Sum test. B.) AADC levels are elevated in early PD participants. Test statistics from Wilcoxen Rank Sum test. C.) CSF AADC levels are associated with clinician-assessed motor symptom severity in Parkinson’s disease participants as measured by the MDS-UPDRS III Off score. D.) CSF AADC levels are associated with clinician-assessed motor symptom severity in Parkinson’s disease participants as measured by the MDS-UPDRS III On score. E.) Effect size and significance for relationships in C,D.

While these findings are encouraging, a key challenge to assessing AADC as a biomarker in our 5 cohorts is its link to a primary treatment modality for PD. AADC is an essential component of dopamine and levodopa metabolism. The possibility that elevated AADC levels are driven by dopamine replacement therapy could not be excluded in our current cohort because nearly all PD participants were taking some form of dopamine replacement therapy at the time of blood and CSF collection.

Therefore, we turned to a CSF and plasma proteomics study generated in the Parkinson’s Progression Marker Initiative (PPMI), which is a multicenter international prospective cohort study that recruited HC and treatment naïve, *de novo* PD participants and collected baseline and annual plasma and CSF. Complete study aims and methodology have been published elsewhere^38,39^. From the PPMI cohort, 37 HC and 36 PD had data available using the same Olink proteomics assay. Data from baseline and longitudinal follow-up averaging 4 years in duration was available, for a total of 219 CSF and 219 plasma samples (see Supplementary Table 9 for demographic and clinical characteristics). To our knowledge, our study is the first comprehensive description of the plasma and CSF proteomics in this cohort.

We examined the baseline visit proteomics in PPMI and found that in treatment naïve *de novo* PD participants, AADC was again the most significantly upregulated protein in CSF, but was not a top upregulated protein in plasma (Figure 3A-C, supplementary Tables 10,11). When considering all timepoints, AADC remained the most significantly upregulated protein in both CSF and plasma (Extended data figure 5, Supplementary Tables 12,13). In PPMI, we found that AADC levels were again significantly associated with symptom severity (Figure 3). CSF AADC levels were associated with MDS-UPDRS III Off scores, (β =1.53, p = 0.020) and MDS-UPDRS total symptom score (β = 2.17, p = 0.045) (Figure 3D-F). Further, plasma AADC levels were also associated with MDS-UPDRS III Off scores, (β =2.45, p = 1.67e-4) and MDS-UPDRS total symptom score (β = 4.18, p = 9.90e-5) (Figure 3D-F), as well as global cognition measured by the MoCA (β = −0.43, p = 0.028). This is in contrast to our findings in the Stanford cohorts, where plasma AADC level is not associated with motor symptom severity (extended data figure 6). The reasons for this difference are unclear, but could be related to the fact that the PPMI cohort is much earlier in disease than the Stanford cohorts (mean time since diagnosis = 0.06 ± 0.05 years vs 6.1 ± 4.3 years).

**Figure 3:**
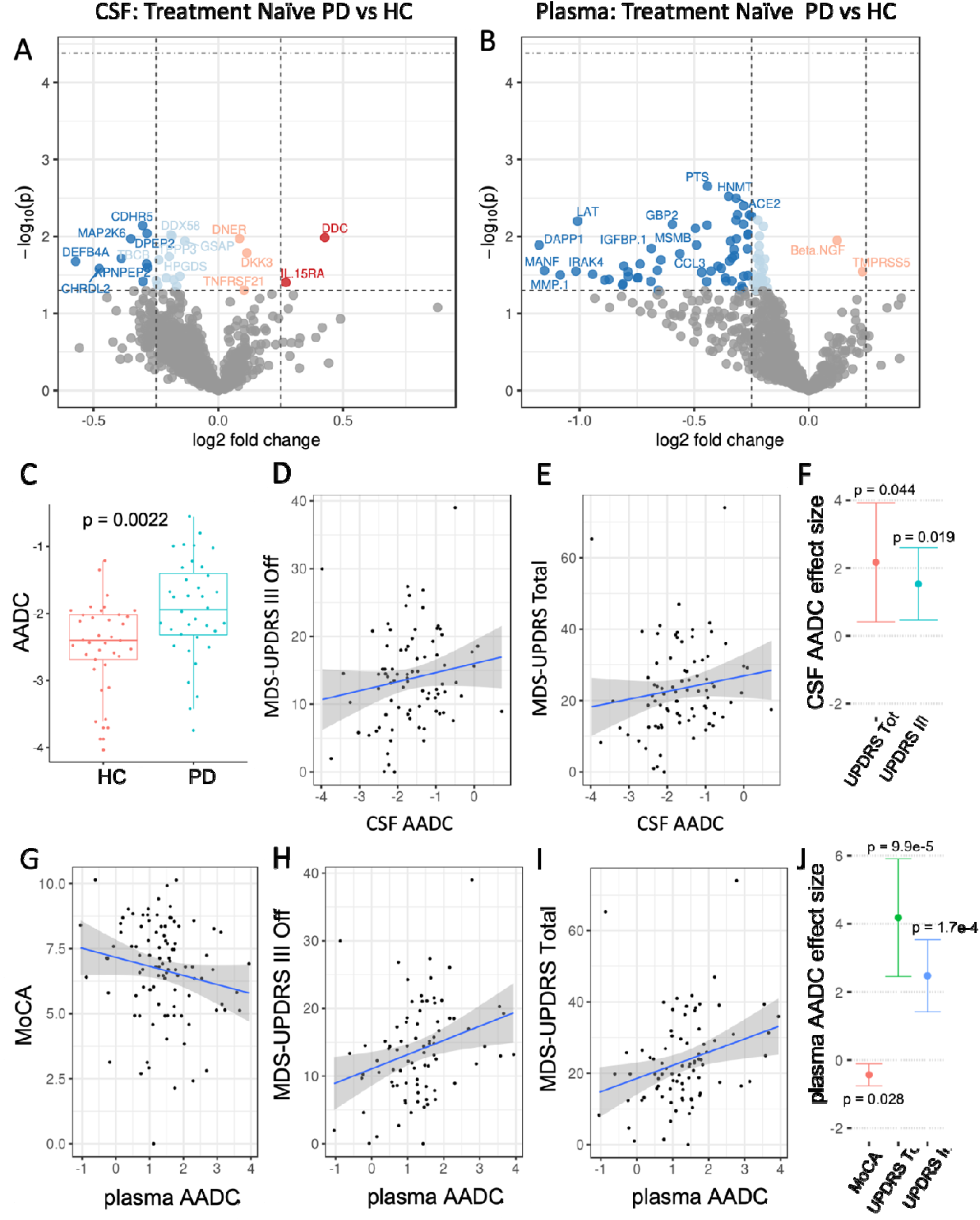
Replication of AADC findings in an independent treatment naive Parkinson’s Disease population. A,B.) CSF (A) and plasma (B) differential expression analysis of treatment naïve Parkinson’s participants at baseline compared to healthy control participants at baseline. Linear modeling controlled for age, sex, and demographics. P values are shown instead of FDR-corrected p values. Due to small sample size of the cohort, no proteins pass FDR multiple testing for proteome-wide significance. C.).) Plot of CSF AADC levels at the baseline visit for treatment naïve PD participants and healthy controls, prior to start of dopamine replacement therapy. D.) CSF AADC level is associated with clinician-assessed motor symptom severity in the PPMI cohort. E.) CSF AADC level is associated with total symptom severity score in the PPMI cohort. F.) Effect size and significance values for models plotted in D,E. G.) plasma AADC levels are negatively associated with global cognition assessed by the MoCA score H.) plasma AADC level is associated with clinician-assessed motor symptom severity in the PPMI cohort. I.) plasma AADC level is associated with total symptom severity score in the PPMI cohort. J.) Effect size and significance values for models plotted in G,H,I

In contrast to these findings on AADC, CSF alpha-synuclein level was not significantly associated with any of these symptoms (Extended data figure 7), suggesting that AADC levels are an earlier and more quantitative correlate of clinical symptoms than CSF alpha-synuclein. Additional trending data that CSF AADC level was associated with time since diagnosis at baseline and time since symptom onset at baseline support the possibility that CSF AADC level in undiagnosed, treatment naïve patients could be a marker of underlying disease progression and dopaminergic neuronal degeneration (Extended data figure 8).

### AADC discriminates PD from AD-s and HC

Given that CSF AADC levels are consistently and uniquely elevated in people with PD, we sought to determine if AADC levels could accurately discriminate PD from HC and AD-s participants. We trained a logistic regression classifier of PD diagnosis against HC and AD-s diagnosis within a subset of the cohorts we measured in this study (PUC/BPD/SCMD), and evaluated its performance in the remaining held out cohorts (ADRC/SAMS) and on the independent PPMI dataset (Figure 4A-E). We found that both CSF and plasma AADC levels were capable of discriminating PD. In the ADRC/SAMS cohort, combining CSF and plasma levels improved classification performance, while in the PPMI cohort, CSF levels alone performed as well as the combination.

**Figure 4.**
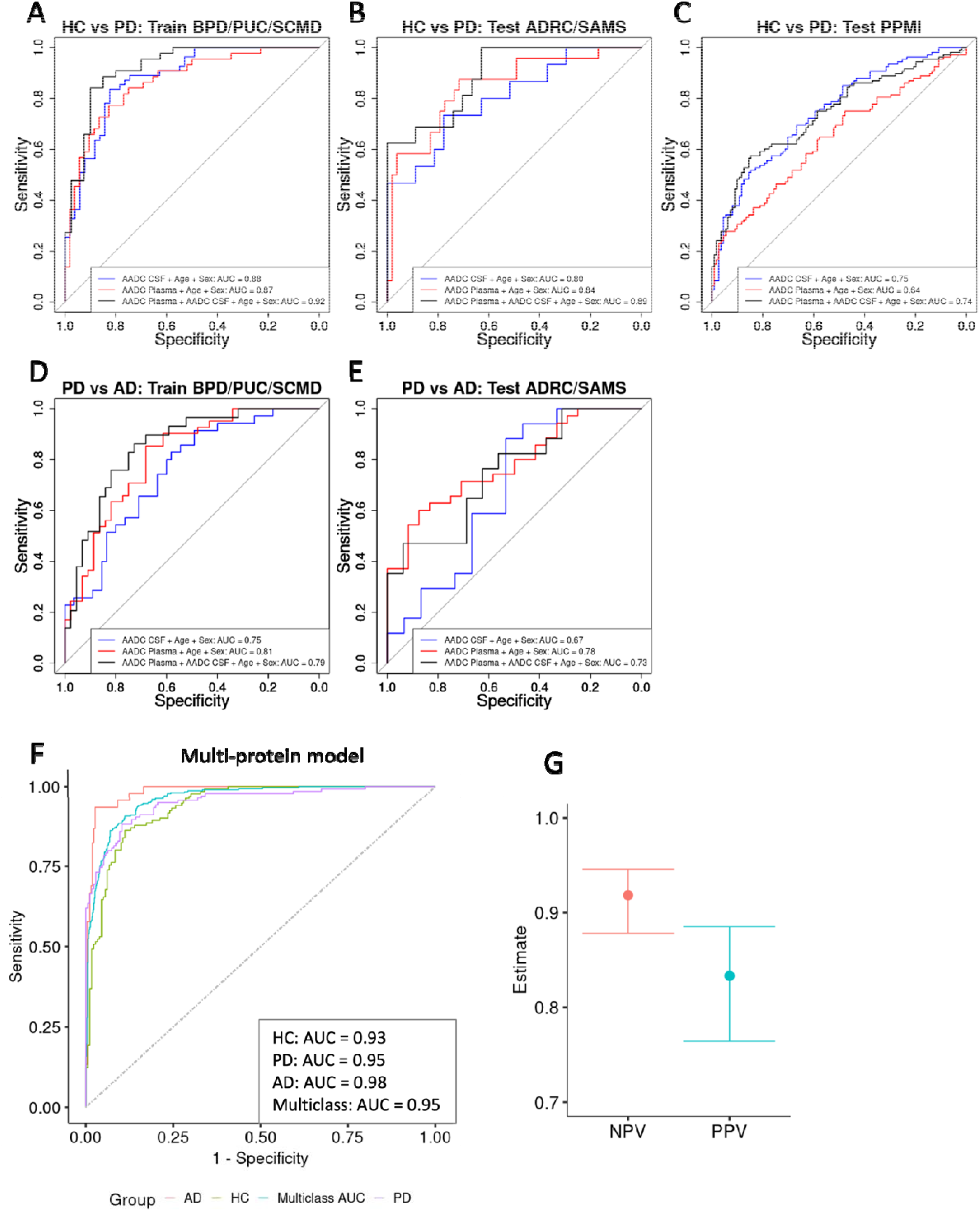
Development and testing of logistic regression classifiers to discriminate PD participants from healthy controls and AD participants using AADC levels in CSF and plasma. Receiver operating characteristic (ROC) sensitivity-specificity curves of classifier performance are plotted. Area Under the Curve (AUC) is a measure of classifier performance, with an AUC of 1 being perfect classification with no false positives (perfect specificity) and no false negatives (perfect specificity). The diagonal line represents the performance of a random guess. Classifier using CSF AADC levels plus age and sex is shown in blue. Classifier using plasma AADC levels is shown in red. Classifier using both CSF and plasma AADC levels is shown in black. Top row A,B,C). ROC curves for the training and testing of the PD vs HC classifier. A.) Performance on PD vs HC in the BPD, PUC, and SCMD cohorts which was used to train the model. CSF AUC = 0.88. Plasma AUC = 0.87. Combined AUC = 0.92. B.) Performance of the PD vs ADHC model in the independent test sets ADRC and SAMS. CSF AUC = 0.80. Plasma AUC = 0.84. Combined AUC = 0.89 C). Performance of the PD vs ADHC model in the independent test set PPMI.. CSF AUC = 0.75. Plasma AUC = 0.64. Combined AUC = 0.74. D,E) ROC curves for the training and testing of the PD vs AD classifier. D.) PD vs AD performance in the training cohorts. CSF AUC = 0.75. Plasma AUC = 0.81. Combined AUC = 0.79 E.) PD vs AD performance in the ADRC/SAMs cohort. CSF AUC = 0.67. Plasma AUC = 0.78. Combined AUC = 0.73. Note there are no AD participants to evaluate in the PPMI so data is not shown. F.) Performance of the 16-protein classification model. Statistics from 2×10 repeated cross-validation shown. Multiclass AUC is calculated by micro-state averaging. G.) Negative and positive predictive values of the multi-protein classifier.

Finally, as a proof of concept for a high-performance molecular biomarker diagnostic tool, we used an iterative approach to generate a minimal biomarker panel that could discriminate people with PD from both HC and AD-s with high accuracy. Starting from a pool of the top 15 differentially expressed proteins in both CSF and plasma for PD compared to HC and for PD compared to AD-s (40 proteins total), we used step-wise forward logistic regression to select a minimal biomarker set with optimal performance using the Akaike Information Criterion (AIC) to select the best performing set. We then trained a penalized multi-class logistic regression model to simultaneously distinguish PD from HC and AD. A model with CSF DDC and 15 additional protein biomarkers plus age and sex had the lowest AIC (Extended data figure 9), and performed with near-perfect classification accuracy across all datasets (Multiclass AUC = 0.95, Figure 4F). We calculated positive and negative predictive value of this classifier and found them to be high (PPV = 0.83, NPV = 0.91, Figure 4G).

## Discussion

There is an urgent need for biomarkers to diagnose PD and to monitor disease severity objectively in clinical trials and clinical practice^7^. In a search for PD-related biomarkers, we discovered CSF AADC (aka DDC) as a potential diagnostic and monitoring biomarker of PD-related neurodegeneration. Our observations show that CSF AADC is specifically and consistently elevated in people with PD, even early in the disease and in treatment naïve individuals. Rather than a global marker of neurodegeneration^40^, our findings suggest AADC might be a more specific marker of monoaminergic neuronal degenerative processes that occur in PD.

A key question for future research is whether elevated CSF AADC is a direct result of neuronal loss or the result of compensatory upregulation in response to neurodegeneration. Both mechanisms would have significant implications for monitoring and therapeutics. Previous studies have suggested that the increasing loss of endogenous AADC in the brain leads to waning treatment responses to dopaminergic medication over time^41–43^. Currently, phase II clinical trials are investigating increasing AADC enzymatic expression via gene therapy as a potential therapeutic intervention for PD motor fluctuations^44,45^. Our data raise the need for further study into the mechanisms of elevated CSF AADC levels. If elevated CSF levels of AADC are due to compensatory upregulation of AADC elsewhere in the brain as a response to nigrostriatal degeneration, this could suggest that the location of AADC in the brain, or other non-dopamine functions of nigrostriatal neurons, play a key role in disease symptoms and progression.

### AADC as a monitoring biomarker of PD

The potential clinical applications of CSF AADC in PD resemble those of serum L-alanine aminotransferase and L-aspartate aminotransferase as biomarkers of hepatocellular damage^46^. Despite their ubiquitous presence in the body, these enzymes are often elevated in hepatic disorders and are implemented as biomarkers of hepatic injury. In parallel, we speculate that increased levels of CSF AADC in people with PD with more severe motor symptoms reflects the underlying pathological progression of the disease, and perhaps even the active degeneration of monoaminergic neurons. This is consistent with the hypothesis that the waning benefit of levodopa therapy over the course of PD is due to declining levels of AADC in surviving dopaminergic neurons. This is also supported by recent gene therapy interventions with an adeno-associated viral vector encoding the DNA for AADC, which show delivery to intact striatal neurons leads to improved patient reported outcomes.^45,47–49^

The primary outcome measure for almost all disease modifying interventions in PD remains the Off-medication UPDRS or MDS-UPDRS. While these are robust measures for severity of motor symptoms^50^11/9/2022 11:15:00 AM, the remarkable placebo effect^51,52^ creates a barrier for exam-based outcome measures in clinical trials. Thus, there is urgency to identify objective measures of nigrostriatal dopaminergic degeneration due to underlying Lewy body pathology^6^. While α-synuclein appeared to be a promising candidate, its application is limited due to the fluctuating CSF levels in PD patients^53^. Here, we found that CSF AADC concentrations are significantly associated with severity of motor symptoms in both treatment naïve patients and those receiving dopamine replacement therapy. The ability to measure severity of the underlying disease regardless of dopamine medications is highly desirable, as this property could both mitigate burdensome Off-medication exams during clinical trials and provide new endpoints for treatment response. We also note the finding that plasma AADC levels showed a relationship with symptom severity in the de novo PD cohort studied here. Given the high potential impact of a minimally invasive monitoring biomarker in early PD, this relationship should be confirmed and investigated in future studies.

### AADC as a specific biomarker of PD

We found that AADC can differentiate PD from neurologically healthy older adults and from adults with Alzheimer’s spectrum disease with good sensitivity and specificity. While it is typically not difficult to differentiate people with PD clinically from people with AD, this analysis lends additional support to the specificity of the underlying disease process measured by AADC and distinguishes it from biomarkers of general brain health such as neurofilament light chain and CSF total Tau^40,54^. Future studies are needed to determine if AADC is also elevated in patients with nigrostriatal dopaminergic degeneration due to non-Lewy body diseases, such as multiple system atrophy or progressive supranuclear palsy, which are challenging to distinguish clinically from PD in early stages of the disease.

In this study, CSF AADC showed similar diagnostic capability to recently developed alpha-synuclein seeding assays (αSyn-SAAs)^55^. In our view, the greatest potential advantage to AADC is its apparent quantitative relationship to disease symptom severity, as αSyn-SAAs have repeatedly shown no relationship to disease severity^55–59^. Other potential advantages include reduced cost, greater robustness, and faster diagnostic times, since αSyn-SAAs are complex assays that take multiple days and specialized equipment to perform. Finally, αSyn-SAA is unlikely to be helpful in LRRK1-PD or Parkin-PD patients, who lack Lewy body pathology despite substantial nigrostriatal degeneration^60^. However, this study is just a proof of concept, and the diagnostic potential of AADC needs to be established in larger cohorts and evaluated with additional methods beyond discovery-focused proteomics. Given that Olink proteomics is an antibody-based platform, we anticipate that the results will translate to other antibody-based detection methods such as ELISA and SIMOA, which could further improve the accuracy and robustness of our results.

### AADC as an early biomarker of PD

Identifying individuals at risk of developing PD could open a window for earlier intervention^61^. Years before the onset of motor symptoms, there is a dramatic loss of dopaminergic, noradrenergic and serotonergic neurons^1,62–65^. We found that CSF AADC concentrations were already elevated at baseline in participants of the PPMI study, all of whom were within two months of diagnosis (average time since diagnosis = 21 days) and had not yet started dopamine replacement therapy. Recent post-mortem studies have suggested that the nigrostriatal pathway terminals degenerate years before substantia nigra neuron death^66^. We hypothesize that AADC may be elevated in the prodromal stages of PD, and future studies should test the prognostic ability of AADC in prodromal individuals.

### Methodological Considerations

This study analyzed discovery proteomics in five Stanford cohorts and one multi-continent cohort from the PPMI. Batch effects between the PPMI and Stanford cohorts affected our ability to train the multi-protein classifier – it was prone to over-fitting which led to decreased test performance between Stanford and PPMI. We believe this is largely due to proteomics batch effects, since training our model in the 2×10 repeated CV paradigm, in which the model learns on a randomly partitioned half of all data (Stanford and PPMI) and its performance is tested on the other half, yielded very high performance. This design limits overfitting better than more traditional CV schemes such as “leave one out” and 10×10 CV. However, 2×10 CV performance is nonetheless likely higher than true out-of-sample performance, so it must be emphasized that this is a proof of concept for high accuracy classification that should be built on in future studies.

While not a multi-site study, we included samples from participants that were recruited independently from five different aging and neurodegeneration cohorts at Stanford. One study strength is that all cohorts include a comprehensive neurological and neuropsychological evaluation as well consensus diagnosis that included a neurologist and neuropsychologist. In addition, all PD participants were evaluated both off and on dopaminergic medications. Further, all five cohorts included at least one longitudinal visit, and three of the cohorts are still being followed either annually, bi-annually or tri-annually (PUC, ADRC, SAMS). Therefore, clinical diagnoses are more accurate than would be from a cross-sectional cohort. For instance, one participant was enrolled as AD-s but at a later visit was found to have clear parkinsonism on exam and met criteria for dementia with Lewy bodies (this participant was excluded from the analysis, see extended data Figure 1). We cannot exclude the possibility that additional PD or AD-s participants had more than one disease contributing to clinical symptoms at the time CSF was collected, but a majority of these participants have agreed to autopsy, which will provide the final pathological diagnoses.

## Conclusion

We have for the first time reported elevated levels of CSF AADC in people with PD and demonstrate that AADC concentrations may be used to differentiate people with PD from healthy older adults and non-nigrostriatal neurodegeneration (AD-s). Notably, AADC is higher in PD participants with more severe motor impairment both on and off dopaminergic medications, and in *de novo* participants with no previous exposure to dopamine replacement therapy. Together, our findings suggest AADC reflects degeneration of dopaminergic neurons in the nigrostriatal pathway as well as other monoaminergic neurons and may serve as an adjunct monitoring and diagnostic biomarker of PD.

## Data Availability

Individual level proteomics data from the present study will be made available upon reasonable request to the authors, contingent on approval for data access via the Stanford Alzheimer's Disease Research Center.

## Acknowledgements

We thank Dr. Jacob Hall, Dr. Veronica Santini, Dr. Sharon Sha and Dr. Laurice Yang for performing lumbar punctures in these cohorts. We thank Michelle Fenesy and Anisa Marshall for assistance with neuropsychological testing and scoring. We thank Jeffrey Bernstein, Nicole Corso, Wanjia Guo, Marc Harrison, Madison Hunt, Manasi Jayakumar, Anna Khazenzon, Celia Litovsky, Natalie Tanner, and Monica Thieu for assistance with SAMS data collection.

We thank participants and family members from the Stanford Alzheimer’s Disease Research Center (ADRC) and the Pacific Udall Center (PUC), and we thank Stanford ADRC and PUC investigators for their contributions to these data. Stanford ADRC investigators are Victor W. Henderson (principal investigator); Administrative Core—Nusha Askari, Katrin I. Andreasson, Victor W. Henderson (leader), Frank M. Longo, Tony Wyss-Coray, and Jerome A. Yesavage; Clinical Core— Carolyn A. Fredericks, Jacob N. Hall, Victor W. Henderson (leader), Kathleen L. Poston, Veronica Rameriz, Allyson C. Rosen, Veronica E. Santini, Sharon J. Sha, Christina Wyss-Coray, Laurice Yang, and Maya V. Yutsis; Biostatistics and Data Management Core—Janet Hwang and Lu Tian (leader); Neuropathology and Biospecimens Core—Donald E. Born, Divya Channappa, Thomas J. Montine (leader), Ahmad Salehi, O. Hannes Vogel, and Tony Wyss-Coray; Outreach, Recruitment, and Education Core—Allyson C. Rosen, Vyjeyanthi S. Periyakoil (leader); Imaging Core—Michael D. Greicius (leader), Elizabeth C. Mormino; Stanford University, Stanford, CA, and the Veterans Affairs Palo Alto Health Care System, Palo Alto, CA.

## Contribution Statement

J.R., K.P., T.W-C., B.L., and P.Z conceptualized the study. J.R. performed all analysis and statistics. J.R. and K.P. wrote the manuscript, which was approved by all authors. B.L. and P.Z. assisted in analysis on early PD. B.L. assisted with binary classifiers. All data was processed and normalized by J.R, B.L., P.Z., and P.M.L. All other authors contributed to data collection in the 5 Stanford cohorts analyzed in this study.

---

**Extended data figure 1:**
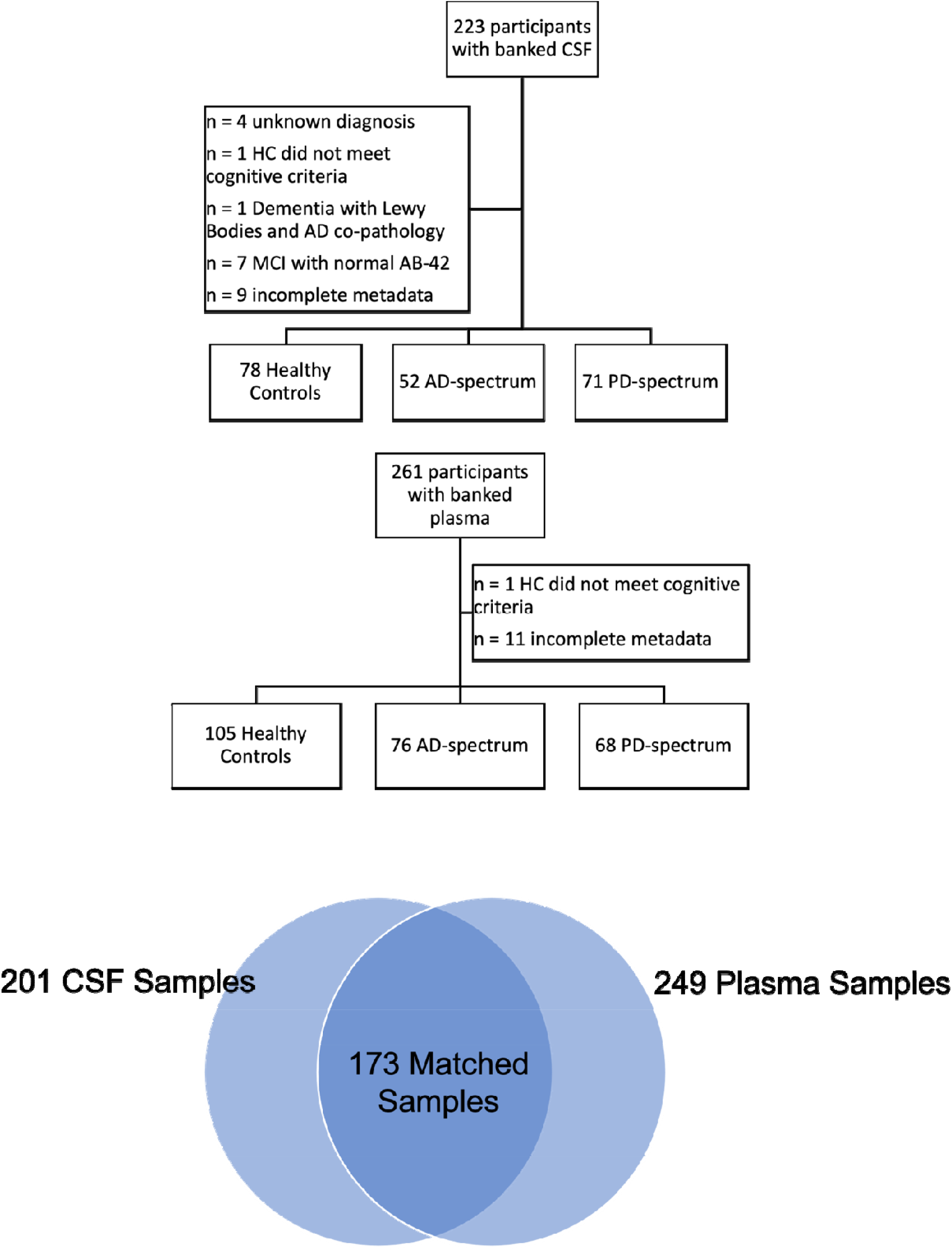
Flowchart of included and excluded participants in the 5 Stanford cohorts analyzed in this study. AD-s, Alzheimer’s Disease spectrum; CSF, Cerebrospinal Fluid; HC, Healthy Controls; PD, Parkinson’s Disease.

**Extended data figure 2.**
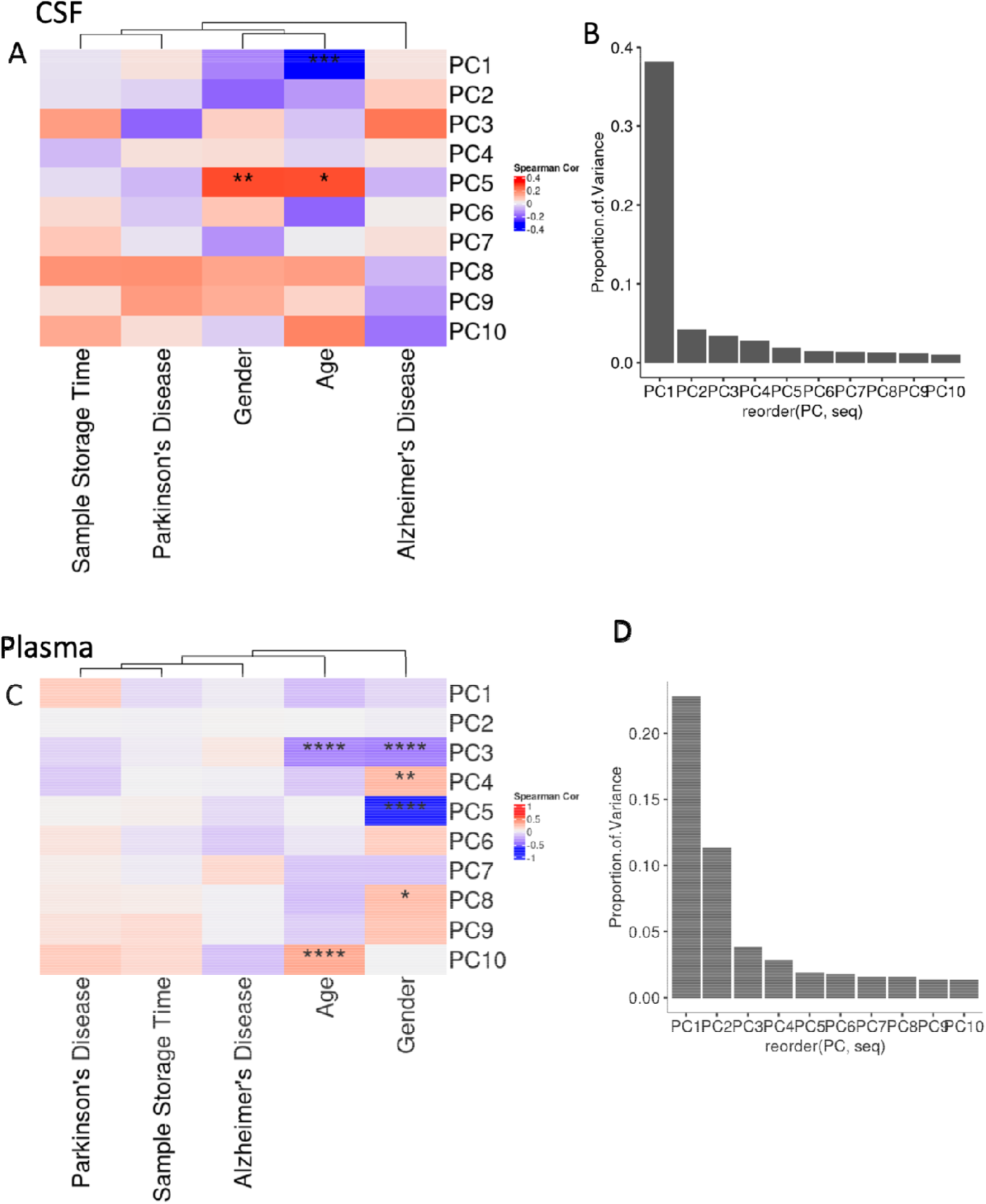
Principal component analysis results for global protein expression in CSF and plasma. A,C) Correlation between top 10 principal components from CSF (A) or plasma (C) protein expression and demographic/disease diagnostic variables of interest. * indicates Bonferroni-adjusted p < 0.05. ** indicates Bonferroni-adjusted p < 0.01. B,D) Scree plots showing the proportion of variance explained by each of the top 10 principal components in CSF (B) and plasma (D).

**Extended data figure 3.**
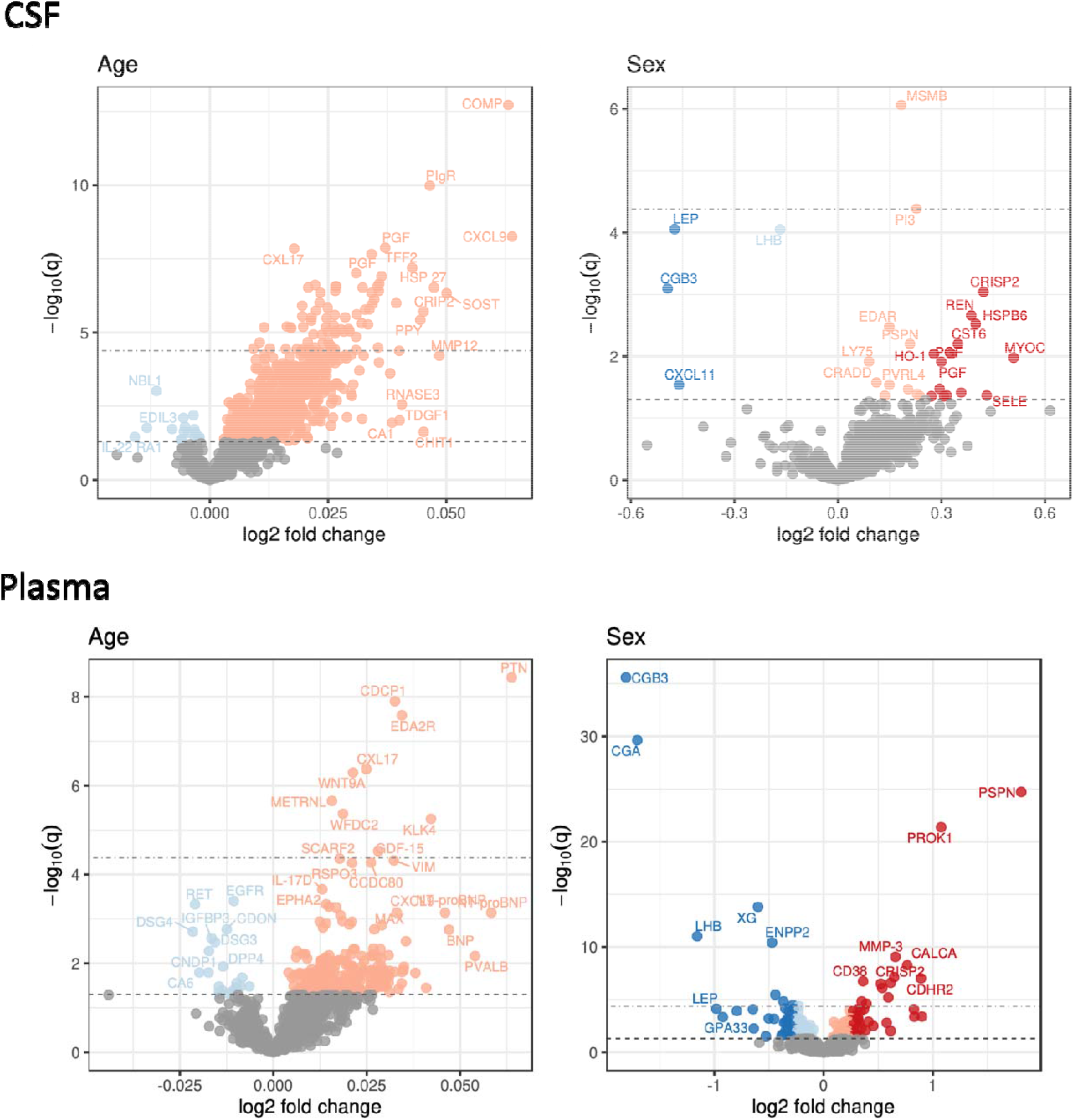
Differential expression of proteomics in the Stanford cohorts for Age and Gender. Lower horizontal dotted line indicates FDR significance threshold, upper horizontal line indicates Bonferroni significance threshold. Dotted vertical lines indicate an arbitrary 0.25 log2 fold change cutoff for differential expression. Significant hits are colored and shaded by cutoff. A,B) CSF expression. C,D) Plasma expression.

**Extended data figure 4.**
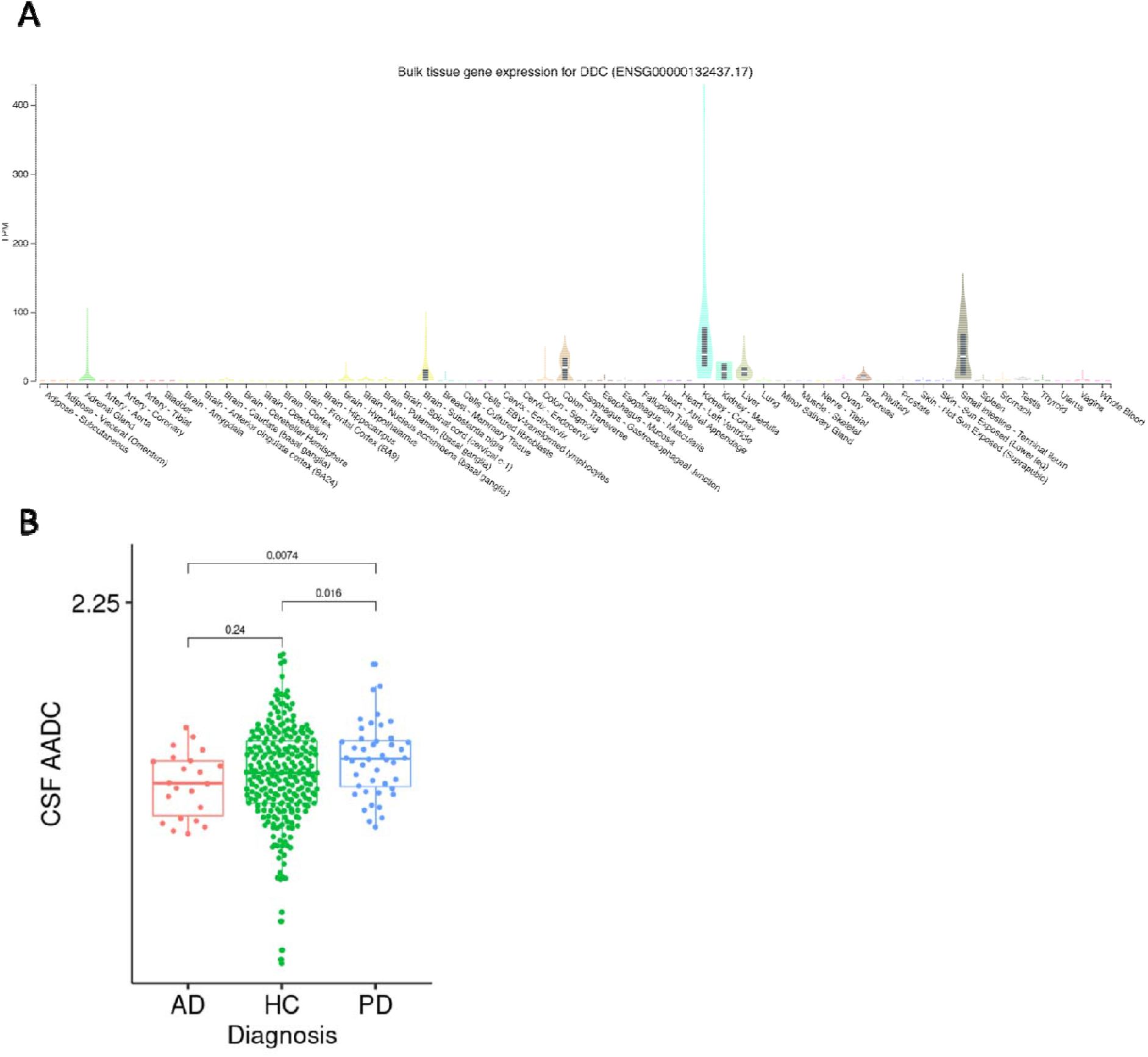
A) Tissue RNA expression level of the DDC gene in the human Genotype Tissue Expression (GTEx) resource. DDC is highly expressed in the substantia nigra of the brain compared to other brain regions.. B) CSF AADC expression measured by the SomaScan proteomics platform confirms levels are specifically elevated in Parkinson’s disease.

**Extended data figure 5.**
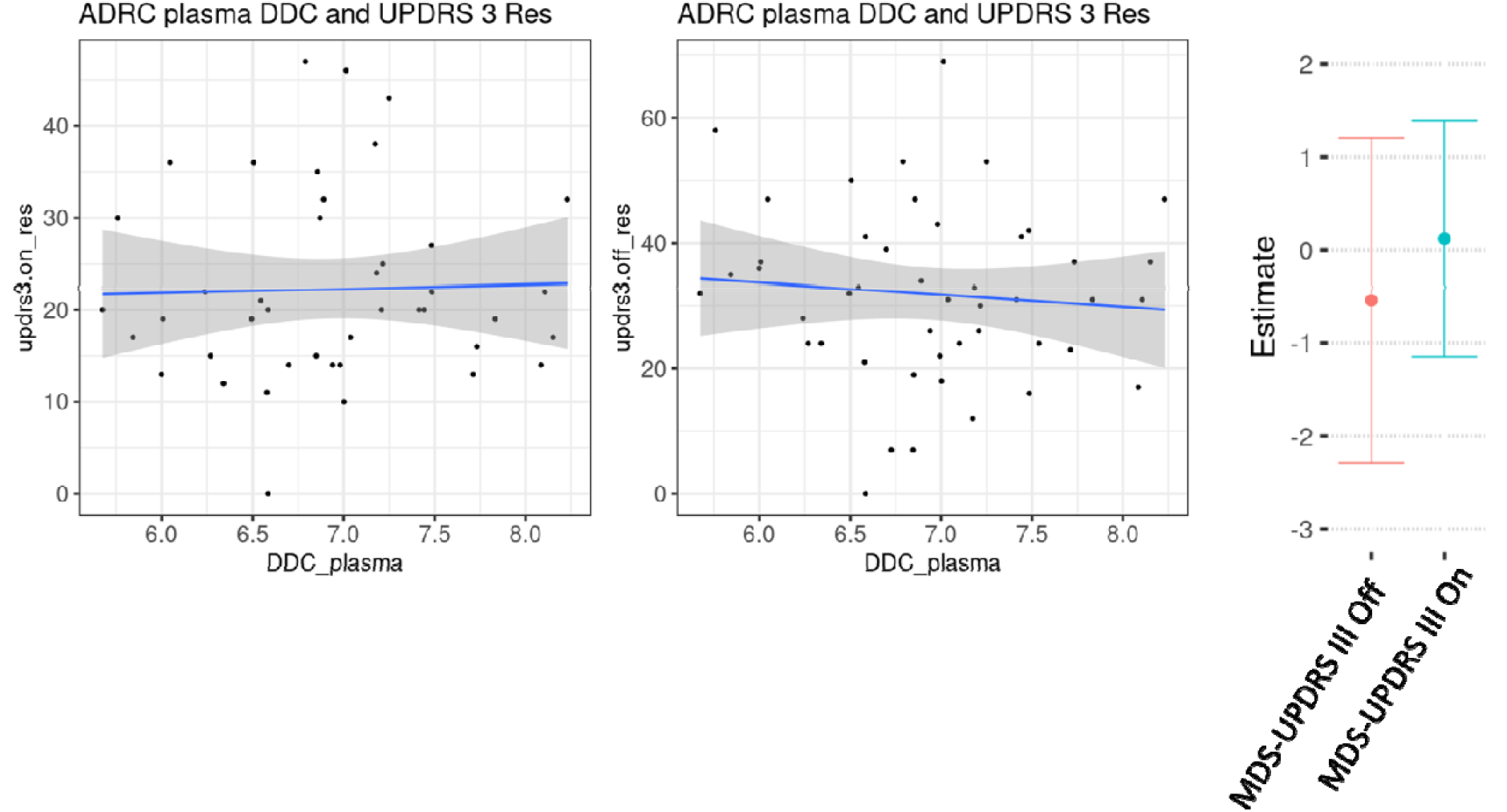
plasma AADC levels are not associated with motor symptom severity in the 5 Stanford cohorts.

**Extended data figure 6.**
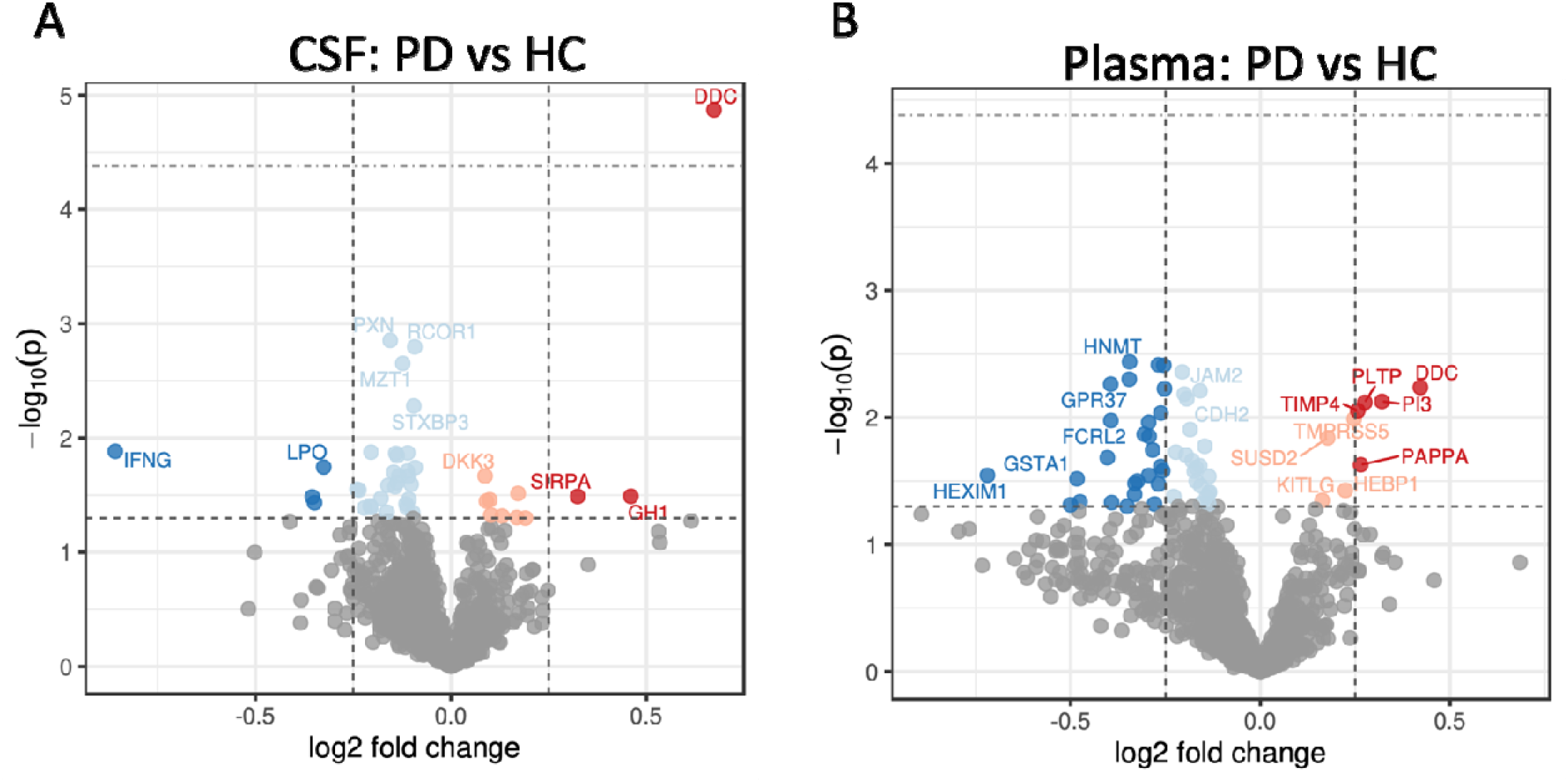
Differential expression analysis of Parkinson’s disease participants in the PPMI cohort using all longitudinal samples. Differential expression is performed with a multi-level mixed effects model controlled for age, sex, demographics, and sample-relatedness. Lower horizontal dotted line indicates FDR significance threshold, upper horizontal line indicates Bonferroni significance threshold. Dotted vertical lines indicate an arbitrary 0.25 log2 fold change cutoff for differential expression. Significant hits are colored and shaded by cutoff. A) DDC (aka AADC) is the only protein that passes proteome-wide Bonferroni significance for differential expression in CSF. P values are shown instead of FDR-corrected p values. Due to small sample size of the cohort, only DDC passes FDR multiple hypothesis testing correction for proteome-wide significance. B) Plasma differential expression as in A.

**Extended data figure 7.**
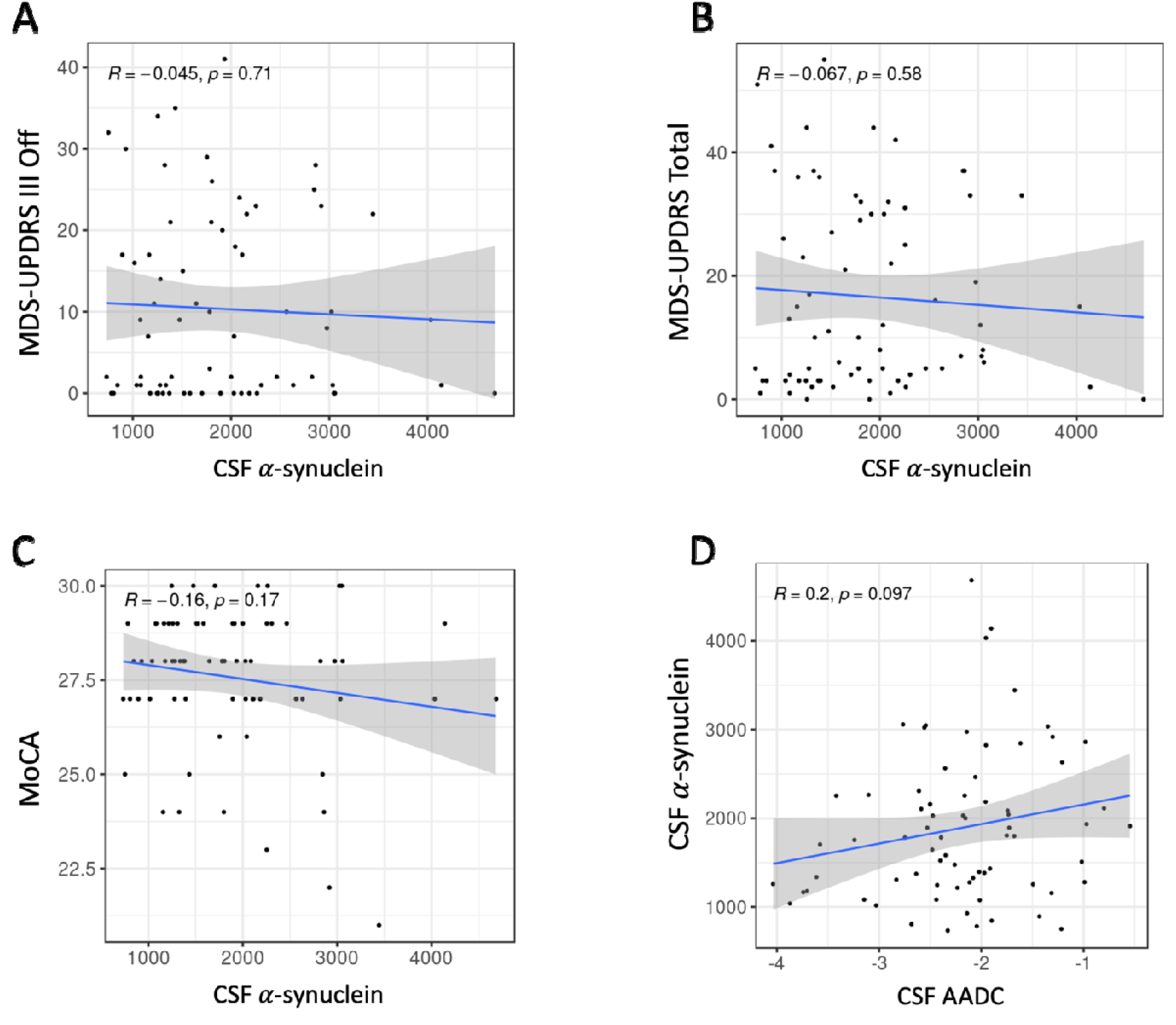
CSF alpha synuclein level is not associated with motor or cognitive symptoms at baseline in the treatment naïve PPMI cohort A) MDS-UPDRS III, clinician-assessed motor symptoms. B) MDS-UPDRS total symptom score. C) MoCA score. D) Correlation between CSF ADC levels at baseline in treatment naïve PPMI cohort and CSF alpha synuclein levels.

**Extended data figure 8.**
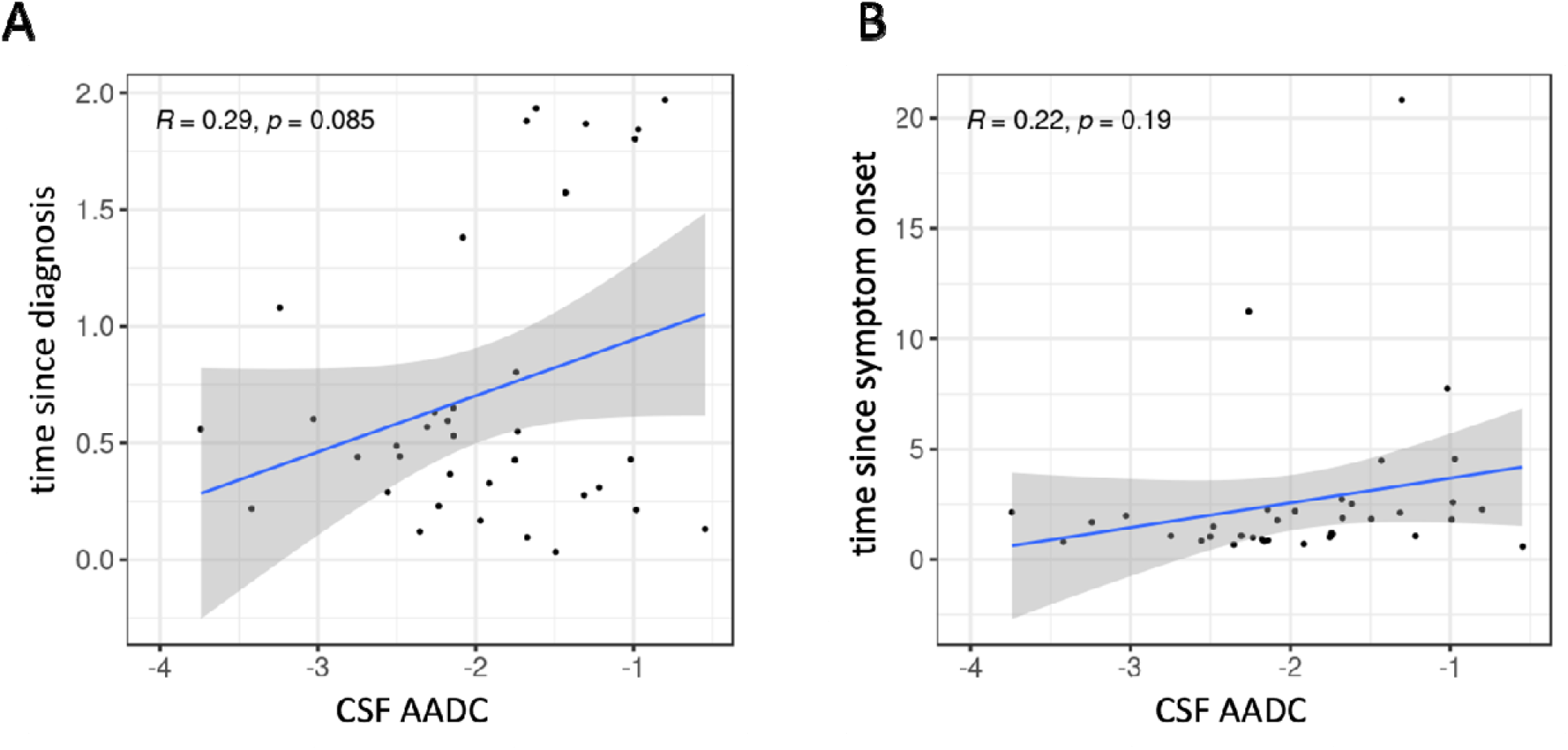
A) Correlation between CSF ADC levels at baseline in treatment naïve PPMI cohort and time since Parkinson’s diagnosis in months. B) Correlation between CSF ADC levels at baseline in treatment naïve PPMI cohort and time since Parkinson’s symptom onset in months.

**Extended data figure 9.**
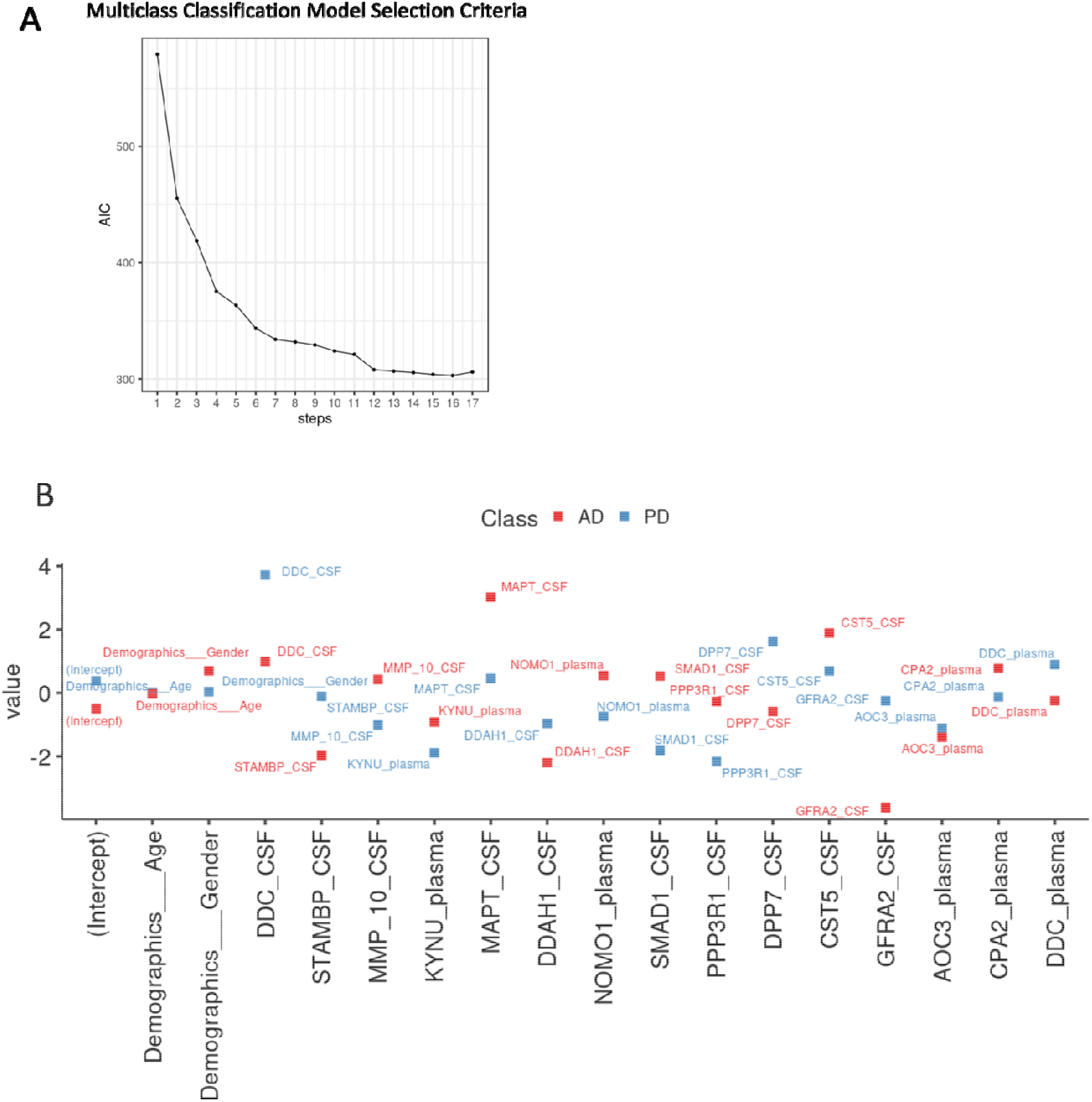
A stepwise multi-class logistic regression was run using the top 15 hits from differential expression of PD vs AD and PD vs HC participants, in both CSF and plasma. A model was built up by iteratively testing the addition of one additional biomarker to previous model (one step) and evaluating the Akaike Information Criteria (AIC) for all possible stepwise combinations. The baseline model included DDC in CSF, age, and gender (step 1). A) The optimal model based on AIC had 15 additional biomarkers, before the AIC began to increase and the model performed sub-optimally. B). Feature weights for the combined biomarker PD classification model trained with L2 penalized logistic regression. Proteins are shown in order of model inclusion using stepwise forward logistic regression.

